# Pitfalls in Time-to-Event Analysis of Registry Data: A Tutorial based on Simulated and Real Cases

**DOI:** 10.1101/2023.12.21.23300363

**Authors:** Mickaël Alligon, Nizar Mahlaoui, Olivier Bouaziz

**Affiliations:** French National Reference Center for Primary Immune Deficiencies (CEREDIH), Necker Enfants Malades University Hospital, Assistance Publique-Hôpitaux de Paris (APHP), Paris, France; Immuno-Haematology and Rheumatology Unit, Necker Enfants Malades University Hospital, Assistance Publique-Hôpitaux de Paris (APHP), Paris, France; Université Paris Cité, CNRS, MAP5, F-75006 Paris, France

**Keywords:** registries, survival analysis, left truncation, competing risks, recurrent events, time-to-event analysis, right censoring, primary immunodeficiencies, rare diseases

## Abstract

Survival analysis (also referred to as time-to-event analysis) is the study of the time elapsed from a starting date to some event of interest. In practice, these analyses can be challenging and, if methodological errors are to be avoided, require the application of appropriate techniques. By using simulations and real-life data based on the French national registry of patients with primary immunodeficiencies (CEREDIH), we sought to highlight the basic elements that need to be handled correctly when performing the initial steps in a survival analysis. We focused on right censoring, left truncation, competing risks, and recurrent events. Our simulations show that ignoring these aspects induces a bias in the results; we then explain how to analyze the data correctly in these situations. Rare disease registries are extremely valuable in medical research. We discuss the application of appropriate methods for the analysis of time-to-event from the CEREDIH registry. The objective of this tutorial article is to provide clinicians and healthcare professionals with better knowledge of the issues facing them when analyzing time-to-event data.

**Key messages box:**

- When comparing naïve approaches and the proper methodology, we show that:
- Not considering right censoring leads to underestimation of survival
- Not considering left truncation leads to overestimation of survival
- Treating competing risks as right-censoring leads to overestimation of survival
- Appropriate recurrent event methods allow to study all events for each patient and not only account for the first event.

## 1. INTRODUCTION

The collection of patient data in academic- and/or industry-led registries is one of the key elements of medical and translational research. The advent of many disease registries (including registries for rare diseases) in the early 2000s helped to improve our knowledge of disease occurrence (incidence and prevalence being the key epidemiological factors most frequently assessed), the natural history of those diseases, and the effectiveness and safety of various procedures and therapies (such as stem cell therapy) at the national and international levels (1,2). Furthermore, registry data is of value in (i) designing national or international orphan drug trials, (ii) standardizing patient management, and thus (iii) improving the patients’ health-related outcomes and quality of life (3).

In France, the creation of a series of five-year national rare disease plans and national reference centers for rare diseases in 2004 prompted the creation of registries for single diseases or groups of diseases. The CEREDIH French national reference center for children and adult patients with primary immunodeficiencies (PIDs) created France’s first national registry for these conditions. The registry complied with the official criteria: the continuous, exhaustive registration of cases (defined as a condition, disease, health issue or healthcare procedure such as surgery, hematopoietic stem cell transplantation [HSCT], etc.) in a defined geographical area by a team of trained professionals (4,5).

PIDs constitute a large, heterogeneous group of more than 500 mostly inherited diseases that expose patients to a greater risk of infections, severe allergies, autoimmune/inflammatory manifestations, and/or malignancies (6,7). Furthermore, PIDs can be classified as deficiencies of the adaptive immune system (subdivided into T-cell deficiencies and B-cell deficiencies) and deficiencies of the innate immune system. The T-cell deficiency group includes severe combined immunodeficiencies (SCIDs, also known as “boy-in-a-bubble diseases”) and other combined immunodeficiencies (CID). The B-cell deficiency group can be subdivided into common variable immunodeficiencies (CVIDs) and hypogammaglobulinemias (also referred to as “non-CVIDs”, which also include agammaglobulinemia).

Since the CEREDIH registry’s inception in 2005, we have sought to include all patients diagnosed with a PID in France (8,9). As of June 2^nd^, 2022, more than 8,500 patients had been registered. 1,563 of these patients are now deceased.

CEREDIH uses the European Society for ImmunoDeficiencies platform to enter data. All European Society for ImmunoDeficiencies registry documenting centers share a common dataset, and CEREDIH has a complementary, specific dataset. Overall, the collected data encompass several medical variables recorded at one or more timepoints in the patient’s life: the symptoms that led to the diagnosis of PID, the main PID-related clinical manifestations (malignancies, autoimmune/inflammatory manifestations, allergies, and infections), the main PID-related therapies (mainstay therapies like immunoglobulin replacement therapy and curative therapies like HSCT, thymus transplant, and gene therapy), and the cause of death. After inclusion, all the patient files are updated every two years or more frequently. Since the dates of these main events are recorded, it is possible to construct time-to-event variables for a given event.

Alongside data completeness, data quality is essential at all stages: at registration and through follow-up documentations as well as through the implementation of relevant and efficient quality control and data management procedures. Furthermore, the entry of multiple time points per patient ensures that information is as up to date as possible and (ii) the quality of the indicators produced by the statistical analyses (especially survival data indicators) is as high.

Underfunding is a concern because it leads to issues of registry sustainability, which include (but are not limited to) understaffing, impairing collaboration with statisticians who have relevant expertise in this field. As a result, some studies may include a suboptimal or even biased statistical methodology, which in turn can lead to incorrect results. One of the primary roles of a registry is to highlight overall trends and relationships in data (10). Research groups can then use specific methods to validate or reject medical hypotheses (e.g. with regard to disease mechanisms, survival, covariates leading to one or more comorbidities of interest, etc.). Therefore, the use of incorrect statistical methods that do not consider potential bias in the data might lead to unreliable estimations and harmful medical decisions. Improving patient management is one of the main goals of patient registries and involves the analysis of time-related data. In the field of health, survival analyses are among those that suffer the most from statistical bias; this is primarily due to the use of inappropriate approaches that do not consider censoring.

Famous examples of incorrect statistical analysis often involve immortal bias. One study (11) found that Academy-Award–winning actors and actresses lived almost 4 years longer than their less successful peers. However, a subsequent reanalysis of the study data failed to find a significant difference in survival between the winners and non-winners and showed that the first analysis suffered from immortality bias: the “*winners had to survive long enough to win”, while “performers who did not win had no minimum survival requirement, and some died before some winners had won, that is, before some “longevity contests” could begin*.” Queen Elisabeth II even joked about immortality bias during her 80^th^ birthday celebration in 2006: “*As Groucho Marx once said, ‘Getting older is no problem. You just have to live long enough*.’”. However, the issue can be more serious when it affects medical research. Using skin cancer as a marker of sun exposure, researchers had concluded that “*having a diagnosis of skin cancer was associated with less myocardial infarction, less hip fracture in those below age 90 years and less death from any cause*.” (12) Following this study, two other researchers – both specialists in the analysis of time-to-event data – pointed out the presence of immortality bias in the first analysis: “*in order to get a skin cancer diagnosis, and thus become a member of the skin cancer group, it is at least necessary to survive until age of diagnosis. For those in the skin cancer group it is impossible to die until the age of diagnosis of the cancer, the so-called immortal person-time*.” Another pitfall pertaining to the study of time-to-event data involves competing risks: if the event of interest is non-lethal (such as disease relapse, an infection, or the occurrence of cancer) and death can also occur, the latter must be treated as a competing event (i.e. an event that precludes the occurrence of the event of interest). A common mistake then consists in treating death as censoring, which amounts to assuming that deceased patients are still at risk of experiencing the event of interest. For example, researchers have compared the risk of relapse among HSCT recipients, using the European Group for Blood and Marrow Transplantation (EBMT) dataset (13). They reported that treating death as a censoring variable resulted in a significant overestimation of the probability of relapse: the estimated 5-year probability of relapse was 0.515 in the flawed analysis and 0.316 when death was correctly taken into account as a competing risk.

Here, we describe the classical methods used to deal with right-censoring, left truncation, competing events, and recurrent events. We first apply a simulation-based approach and then refer to CEREDIH registry data. Our objective is to make clinicians and healthcare professionals more aware of the issues facing them in analyses of time-to-event data.

All analyses were conducted with R software and its *{survival}* library. All the codes and a randomized version of the CEREDIH dataset are available on GitHub (https://github.com/Malligon/Pitfalls-in-Time-to-Event-Analysis-for-Registry-Data).

## 2. HOW TO PLAN YOUR SURVIVAL ANALYSIS CAREFULLY

Survival analysis (also called time-to-event analysis) is the study of the time elapsed from a starting date to an event of interest.

Firstly, it is important to precisely define the event of interest, the time scale, the study entry point, and the risk set. The event of interest can be death, recovery, occurrence of a disease, relapse or any medically relevant event. The time scale refers to the time unit used (usually years or months). Study entry is the starting point of the study (birth, treatment initiation, enrolment, etc.). If, for example, a study is designed to analyze survival (in days) after treatment, study entry will be the time at which the patient took his/her treatment, and the time scale will be days. Lastly, the risk set is defined as the pool of patients at risk of experiencing the event of interest. A patient is included in the risk set at a specific time if he/she can experience the event of interest at that time; this means particularly that a patient can enter and leave the risk set at any time.

A classical phenomenon in time-to-event analysis is the presence of incomplete data. This can be caused by right-censoring, left truncation, or both. These data might also include recurrent and/or competing events. Failure to take these concepts into account may lead to incorrect estimations and misleading conclusions. Below, we present these four statistical concepts and we explain how they can be handled by properly adjusting the risk set in each case.

For some individuals, the exact time of occurrence of the event of interest is not known; instead, an earlier time is observed, and it is only known that the event of interest will occur after this observed time. This is ***<u>right-censoring</u>***, which is classically taken into account with the Kaplan-Meier estimator.

***<u>Left truncation</u>*** is a phenomenon that often occurs in time-to-event analysis in which individuals are followed up only from a time after study entry (called the *truncation time*) and not from study entry onwards. In such a case, individuals are observed conditionally on having not experienced the event of interest before the truncation time. In order to avoid biased estimates, those data need to be appropriately taken into account by modifying the risk set in the Kaplan-Meier estimator.

***<u>Competing risks</u>*** methods are involved in a situation that occurs when another event may preclude the observation of the event of interest. This is typically the case when the competing risk is death and the event of interest is the occurrence of a disease, remission, the onset of cancer, etc… While censored data indicates that the true event of interest will occur after the censoring time, the true event of interest can no longer occur after a competing risk. A common error consists in treating competing events as censored data in the calculation of the survival function of the event of interest. This leads to overestimation of the distribution of the event time. The correct approach consists in estimating the cumulative incidence function (CIF), using specific methods.

***<u>Recurrent event</u>*** data occur when an individual can experience the event of interest several times during his/her lifetime. This can happen for the study of recurrent infections, hospital admissions, cancer relapses, etc… Furthermore, recurrent event data often include a competing event (referred to as the terminal event), which is typically death. For these data, a different quantity may be of interest, such as the expected cumulative number of recurrent events experienced by a patient up to a given time point.

### TOOLBOX 1

**How to plan your survival analysis carefully.**

- Focus on data completeness during a chosen time period, rather than focusing on its length.
- Always start by defining the event of interest, the time scale, study entry, and the risk set.
- Assess right-censoring, left truncation, and competing risks in advance and use dedicated methods to analyze those data.
- Consider a recurrent event analysis if patients can encounter the event of interest more than once during the study.
- In general: always consider what <u>might</u> occur, rather than what has been observed.

## 3. A SIMULATION-BASED APPROACH

### A-Right-censoring

As mentioned above, a time-to-event analysis will usually include right-censored data. Right-censoring can mainly occur for two reasons: (i) the patient has not yet experienced the event of interest by the time the study ends, or (ii) the patient is lost to follow-up during the study period (i.e. drop-out). Because such censored times are smaller than the true event times, treating the censored observations as completely observed data will underestimate the distribution of the true event times. In contrast, larger and smaller times of interests will respectively tend to be more or less subject to right-censoring. As a result, keeping only the uncensored observations will result in underestimation of the true event time distribution. These two naïve approaches (treating censored data as completely observed, or removing censored data) show that dedicated methods (namely the Kaplan-Meier estimator, in the case of right-censoring) are needed in this context.

When dealing with time-to-event data with right-censoring, the observations for an individual consist of two variables: the observed time and the censoring status (or censoring indicator). The latter variable is binary and indicates whether the observed time is the time of interest or the censored time - a time that is known to be smaller than the time of interest. In order to simulate these types of data for each individual, one needs to: (i) simulate the time of interest; (ii) simulate a censoring time; (iii) measure the minimum between the two times, which is the observed time; and (iv) determine whether the time of interest is smaller than the censoring; if so, the censoring status is equal to 1; if not, the censoring status is equal to 0. In the ***<u>Appendix</u>***, we present a simple code that simulates these data as described above. We chose a Weibull distribution for the true event time (with a shape of 2 and a scale of 30) and a uniform distribution over the interval [0;85] for the censoring variable. On average, this choice of parameters will result in 31.24% of censored data. A sample of size 1000 is simulated. It should be noted that a seed was arbitrarily chosen, so that the data can be reproduced easily (see the *Appendix* for more details). **Table 1** shows the first 10 individuals simulated using this code.

**Table 1:**
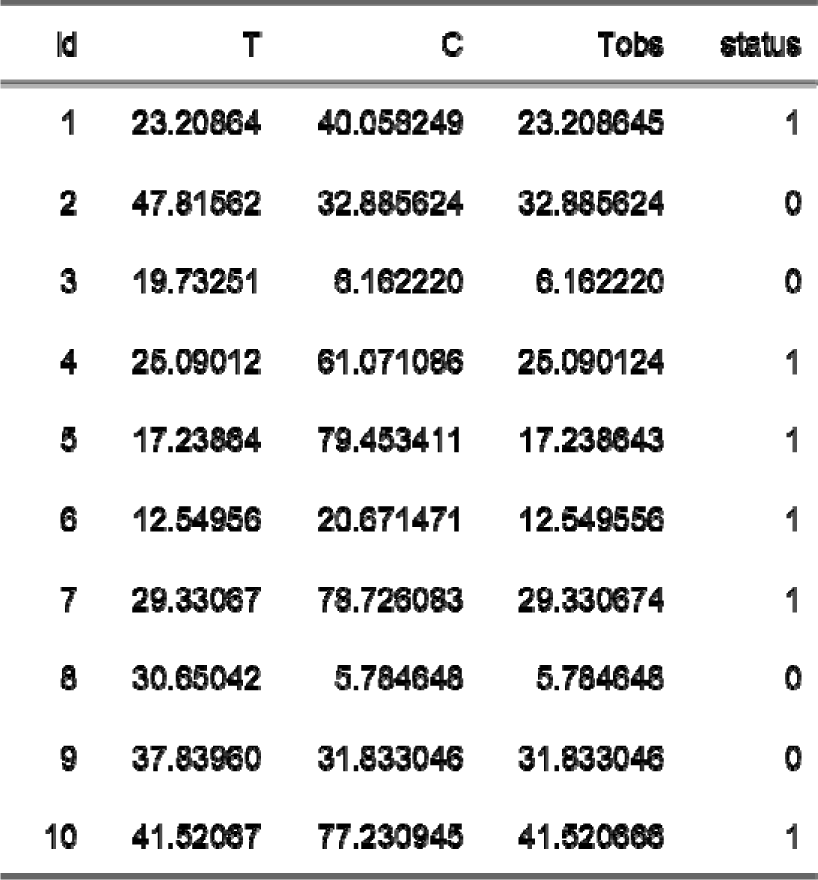
the first 10 individuals simulated.

**<u>In the absence of covariates, the standard quantity of interest in survival analysis</u>** is the **<u>survival function</u>** which represents the probability that the event of interest has not yet occurred at any given time point. The survival function is classically estimated using the Kaplan-Meier estimator (14). Starting from the grid of time points t_1_, …, t_K_ consisting of all the uncensored times, the estimator is computed recursively. For a given t_k_, the Kaplan-Meier estimator is equal to its value at the previous time point t_(k-1)_ times (1-d_k_/R_k_), where d_k_ is the number of uncensored events that occurred at time t_k_, and R_k_ is the number of “at risk” individuals at time t_k_ (defined as the number of individuals that have not yet experienced the event of interest or who have not yet been censored). In this formula, the estimator is initialized at the value 1 for time equal to 0. In survival analysis, the notion of individuals being “at risk” at a given time is essential. It is only through this risk set that censoring is accounted for, and it is crucial that this set does not include periods of time during which the event of interest cannot occur. The d_k_/R_k_ ratio is called the hazard rate or hazard risk estimator and represents the estimation for the risk of experiencing the event of interest at time t_k_, given that this event has not yet occurred. By using the *survfit* function in the R package ‘survival’, one can compute the Kaplan-Meier estimator for the previously generated dataset. This estimator can be compared with the naïve approach described in the second section, which consisting in removing the censored observations and computing 1 minus the empirical distribution function for this subsample (**Figure 1**; see the *Appendix* for more details). Clearly, the Kaplan-Meier estimator produces a very accurate estimation, whereas the naïve approach gives a biased estimation. As expected, the survival function is underestimated when censoring is ignored. For example, the true quantiles (Q_1_, Q_2_ and Q_3_) for the variable of interest are 16.14, 25 and 35.37, respectively. The three quantiles estimated from the Kaplan-Meier estimator are 16.12, 25.09 and 35.95, respectively, while those given by the naïve estimator are 13.95, 22.06 and 30.42, respectively. By way of an example, one can consider a study in which the event of interest is death: including only observed deaths in the analysis (and thus ignoring censored data due to the end of study or drop-out) will result in underestimation of the survival function. This is because at all time points, the risk set needs to include the censored observations that have not yet occurred. Whereas the number of observed events of interest is the same (d_k_ in the Kaplan-Meier estimator), the number of individuals at risk (R_k_ in the Kaplan-Meier estimator) should be increased.

#### TOOLBOX 2

**Right censoring**

- Right censoring is extremely common in time-to-event analyses.
- The Kaplan-Meier method is the standard approach for estimating the survival function for right censored data.
- Ignoring right censoring leads to underestimation of the survival function.
- When observations are censored, the event of interest will happen at a later, non-observed time.

**Figure 1:**
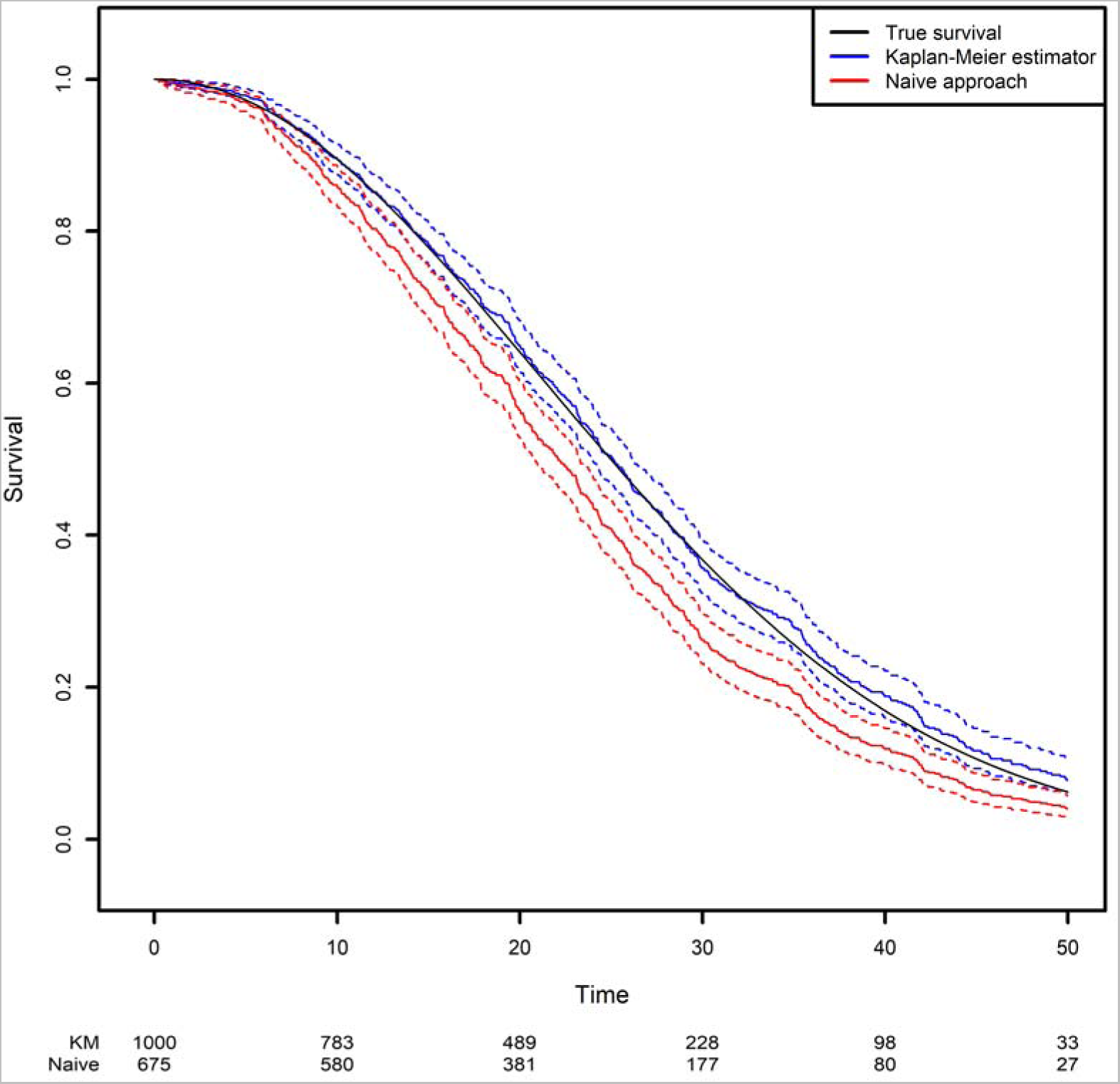
Comparison of the estimated survival function using the naïve approach (in which censored observations are removed) and the Kaplan-Meier estimator.

### B. Left truncation

Another frequent phenomenon in time-to-event analysis is left truncation (delayed entry), when individuals are followed from a later time (the truncation time) and not from the starting point. In such a case, individuals are observed conditionally on having not yet experienced the event of interest before the truncation time. Again, in order to avoid biased estimates, those data need to be taken into account appropriately by modifying the risk set in the Kaplan-Meier estimator. While right-censoring is often correctly taken into account in the analysis of time-to-event data, left truncation is more difficult to apprehend and is therefore sometimes overlooked. Not taking into account left truncation results in an immortality bias because individuals are considered to be at risk before the truncation time but cannot die – if death is the event of interest – before the truncation time. This is typically the case when the time scale is age since, very often, patients cannot be followed up from birth. In that case, it is important to take into account the data observation scheme: depending on the study, an individual will start to be followed up at the time of diagnosis, at the date when the treatment started, or at some other time. Individuals having experienced the event before they started to be followed up will never be observed. If the time scale is age and the patient enters the study at a specific time, then he/she should not be part of the risk calculation for earlier times.

Left truncation can be easily taken into account by modifying the risk set in the Kaplan-Meier estimator. At a given time point, an individual should be in the risk set if he/she (i) has not yet experienced the event, (ii) has not yet been censored, and (iii) the truncation time occurred earlier than the time point.

It is important to stress that ignoring left truncation would result in overestimation of the survival function because the risk set would be too large at time points where all patients have not yet entered the study. Since the Kaplan-Meier estimator is computed in a recursive way, this bias for initial time points will have an impact on all later times, and this incorrect survival function will be overestimated.

Using the same simulation scheme as before, we generated a truncation variable with a uniform distribution over the interval [0;50]. As a result, 42.4% of the observations are not observed because the event of interest occurred before the truncation time. We then estimated the survival function by applying two approaches based on the Kaplan-Meier estimator: the correct one that modifies the risk set according to the truncation variable, and a naïve approach in which left truncation is ignored. Modifying the risk set in the Kaplan-Meier method is easily achieved in the survival library by using the start and stop variables instead of the usual observed time variable. The start and stop variables correspond respectively to the truncation time and the observed time (***Figure 2***; see the ***Appendix*** for more details).

**Figure 2:**
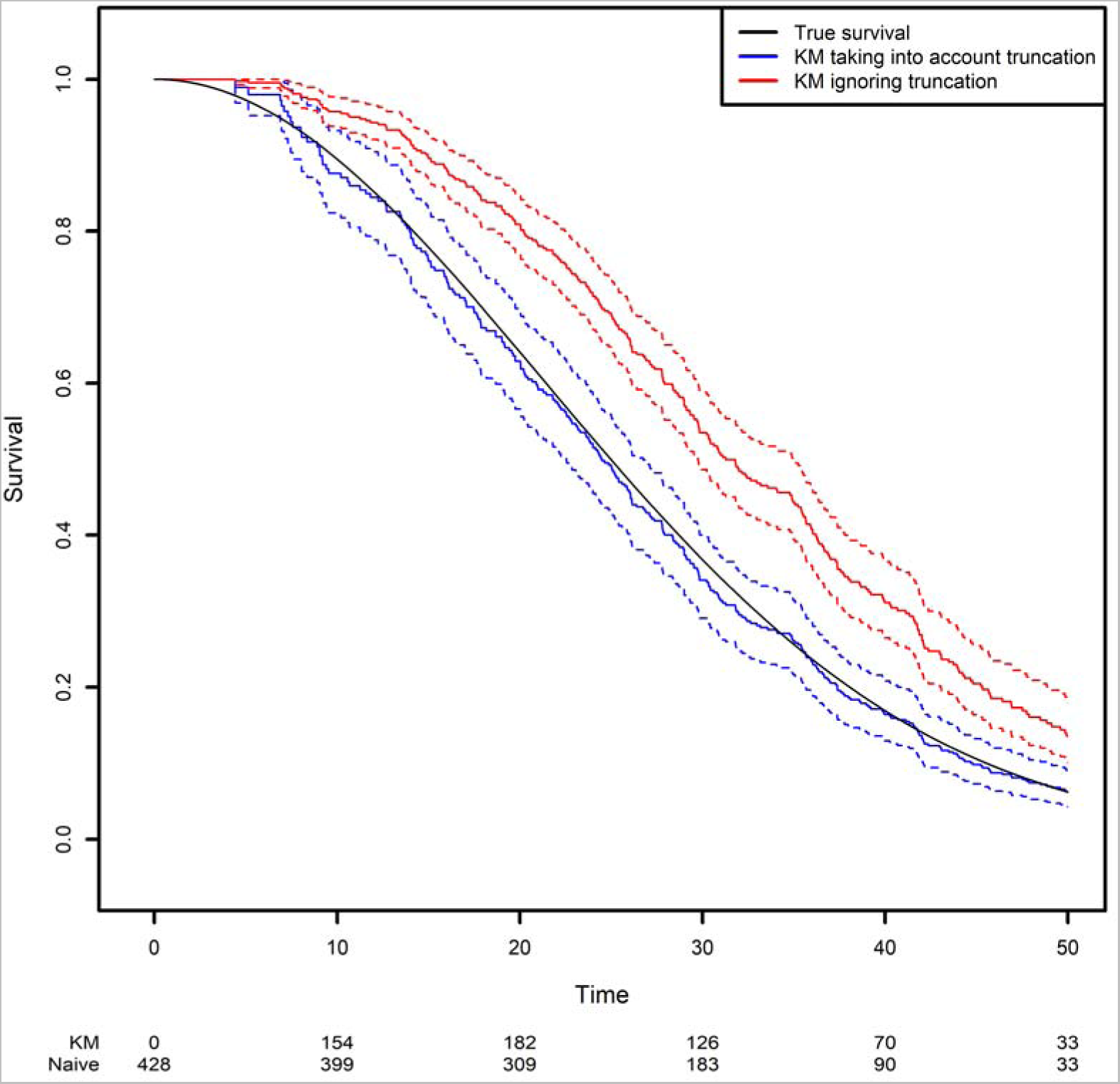
Comparison of the estimated survival function using the Kaplan-Meier estimator that takes left truncation into account and the KM estimator that ignores left truncation.

One can see clearly that the naïve approach overestimates the survival function. For example, the true quantiles of order 0.25, 0.5, 0.75 for the variable of interest are equal to 16.14, 25, and 35.37, respectively. The estimated quantiles from the Kaplan-Meier estimator when left truncation is taken into account are equal to 15.14, 24.44, and 35.43, respectively, while the estimated quantiles from the naive estimator are equal to 22.58, 31.25 and 42.14, respectively.

In summary, it is important to check that a survival analysis’ risk set is well defined. In other words, the researcher should ask him/herself “Is there a period of time during which the individuals cannot experience the event of interest?”. If so, then the risk set needs to be modified accordingly by using the start and stop variables. Similarly, it is important to choose an appropriate time scale for use in practice. Most of the time, this choice will be based on medical considerations. Does it make more sense to study the risk of death on the age time scale? Or should the scale be the time elapsed since treatment allocation? In the second scenario, a wide age range might make it necessary to also adjust for age. In the first scenario, it is very likely that the data will suffer from left truncation. Lastly, it should be noted that left truncation might deteriorate the performance of the Kaplan-Meier estimator when the risk set is too small for short time periods. A small risk set will result in a high hazard rate and a high variance of the hazard rate. Given that errors at early times will have an impact on all future time points, this issue can be problematic. Some other options for managing this problem can be found in the literature (15).

#### TOOLBOX 3

**Left truncation**

- Left truncation is very common in registry analyses - especially when patients are followed up from birth.
- Left truncation is a specific type of immortal time bias.
- The Kaplan-Meier estimator accommodates with left truncation by adjusting the risk set (adding patients or removing them) at a given time point.
- Ignoring left truncation will lead to overestimation of the survival function.
- Early events may impact and bias the survival function if the risk set is too small at early time points.

### C. Competing risks

As mentioned above, competing risks occur when another event may preclude the observation of the event of interest. This is typically the case when the event of interest is not terminal, e.g. the occurrence of an infection or a diagnosis of cancer. Death is then a competing event and if it occurs in the dataset, it must be properly taken into account. A common mistake is to treat death as a censoring variable. The major difference between a competing event and censoring is that the event of interest may occur after the censoring timepoint (even though it is not observed) but will never occur after a competing event. If the competing event is death and the event of interest is cancer, then it is clear that a patient can no longer develop a cancer after he/she had died. The Kaplan-Meier estimator treats censoring as a variable that stops the observation of future events for the patient but includes the information that the event of interest will occur after the censoring variable. Consequently, computing a survival curve using the Kaplan-Meier estimator in a competing risk situation where death is treated as a censoring variable will give a biased estimation. Since dead individuals will remain “at risk” in the computation of the survival function, the estimate will be biased upwards and the survival curve will be overestimated.

Competing events are often not correctly analyzed because they can be treated as a censoring variable when estimating the hazard rate. This is a computational trick that works well because the hazard rate is a quantity defined for an infinitesimally short period of time. In other words, studies that use the Cox model to evaluate the effect of one or more covariates on the event of interest might treat the competing event as a censored variable (16). This approach will provide correct estimates of hazard ratios. However, this practice is no longer appropriate for estimating cumulative quantities, such as the cumulative hazard function or the survival function. Since the Cox model is beyond the scope of this paper, we shall not discuss this issue further.

It should also be noted that the last example when cancer is the event of interest and death is the competing event is more precisely an illness-death situation. Strictly speaking, competing event situations encompass data for which the events of interest are mutually exclusive (17). A typical example is when different causes of death are recorded and analyzed. Again, one cause of death can only occur if the other cause of death has not yet occurred, and this has to be properly taken into account in both scenarios. In the cancer/death example, the death event might be studied simply by computing the Kaplan-Meier estimator because cancer does not preclude the occurrence of death. We nevertheless chose to simplify the presentation by considering this example with cancer and death, because the illness-death model (a particular example of a multistate model (MSM)) is beyond the scope of this article (17). Furthermore, situations in which cancer is of interest and individuals are also at risk of death are frequently encountered in registry data. This will be illustrated below on the CEREDIH dataset.

In the presence of competing risks, the quantity of interest is usually the CIF. For the cancer example, the CIF is simply the probability of experiencing a cancer before any time point. For a given time t it is computed by cumulating for all time points t_k_ occurring before t, the product of the hazard risk for the event of interest (computed as the ratio d_k_/R_k_ at t_k_, where the risk set R_k_ includes individuals that have not yet experienced any of the different types of events and have not yet been censored) and the probability to have “survived” up until time t_k_. This last quantity is basically the Kaplan-Meier estimator for the compound event composed of all the types of event; in other words, it is the Kaplan-Meier estimator where the event of interest is the first event among all competing events). This estimator can be calculated from the survival library by simply considering the status variable as a factor with more than two levels: one level (always the first) for censoring and the other levels for the competing events (18).

Lastly, given that the competing event precludes the occurrence of the other event, it is good practice to always displaythe CIFs of all the competing events as well as the CIF of the quantity of interest (19). This is important because otherwise, the CIF of the quantity of interest might be misleading. A low risk of experiencing an event might simply be due to the fact that the patients are at high risk of experiencing the competing event. Taking again the cancer/death example, individuals might be at a low risk of cancer only because they are at high risk of dying. Another illustrative example (from the CEREDIH dataset) will be given later.

We generated two competing events, along with a censoring variable (**Figure 3**; see the ***Appendix*** for more details). The CIF was calculated in two different ways: the correct way, by considering the other event as a competing risk (as described above), and the naïve approach based on a Kaplan-Meier estimator where the other event is treated as a censoring variable (in the latter case, the curve is obtained by computing one minus the Kaplan-Meier estimator). One can see clearly how important it is to analyze competing risks correctly: the naïve approach clearly overestimates the CIFs. Again, this is because the naïve approach considers individuals to be at risk after they have died, as illustrated by the fact that both curves tend to 1 as time goes to infinity. In contrast, with the correct method, the sum of the two probabilities tends to 1 as time goes to infinity; each individual will experience one (and only one) of the two events with probability one in the future.

**Figure 3:**
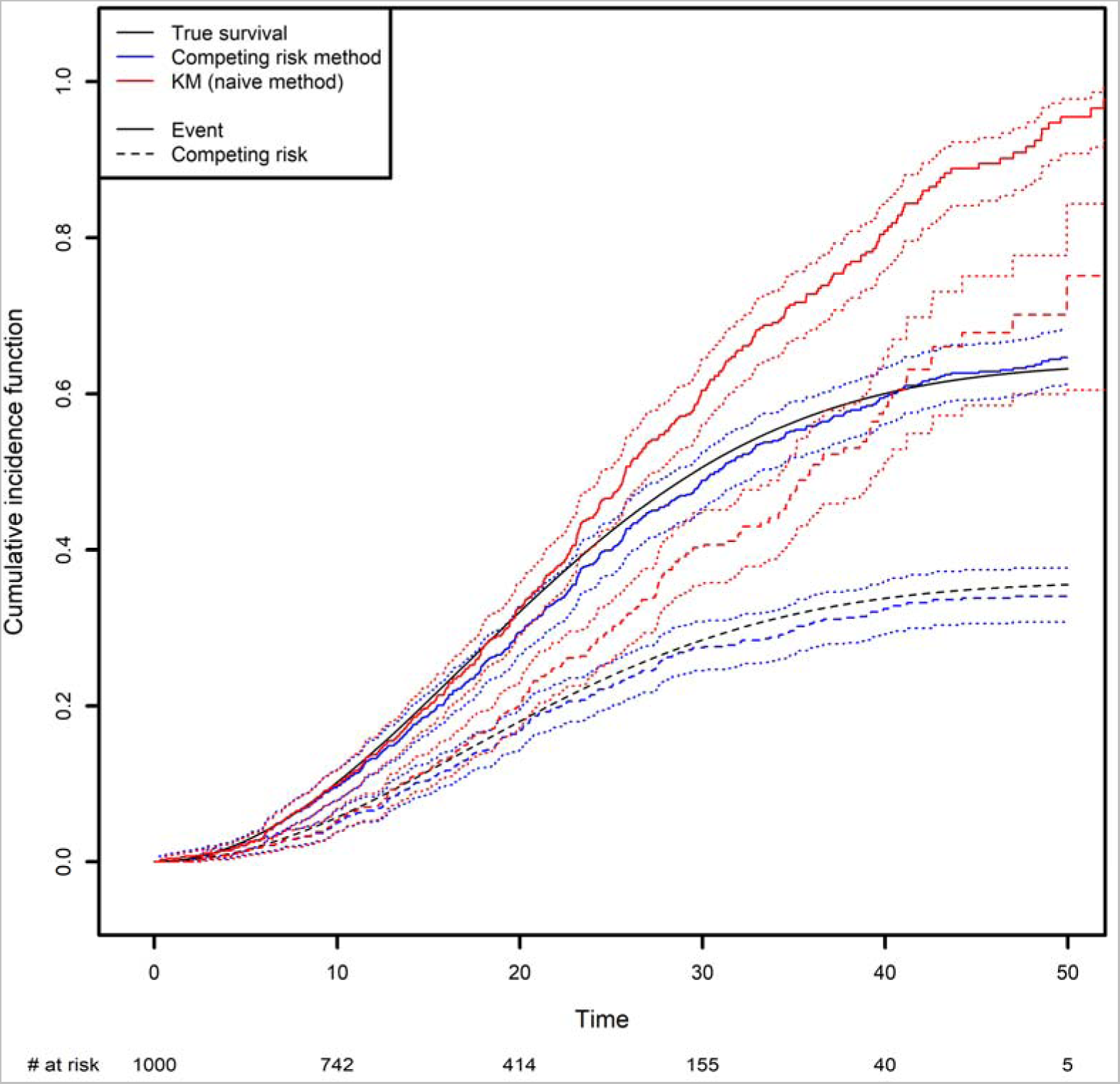
Comparison of the estimated cumulative incidence function using the competing risk method and the Kaplan-Meier estimator, which treats the competing risk as censoring.

In the ***<u>Appendix</u>***, we present some simple code that simulates a competing risk situation. We chose a Weibull distribution for the true event time of interest (shape: 2; scale: 30) and for the competing event (shape: 2; scale: 40). This choice of parameters will result in 24.98% of censoring, 48% of observed events of interest and 27.01% of observed competing events on average. The true curve was implemented based on calculations from the Supplementary Data 1.

The true quantiles of order 0.1, 0.2, 0.3 for the competing event are equal to 13.69, 21.61 and 32.12 respectively. The estimated quantiles from the competing risk method estimator are equal to 14.19, 22.28 and 35.15, respectively, while the estimated quantiles from the naive estimator are equal to 13.57, 19.88 and 25.10, respectively.

#### TOOLBOX 4

**Competing risks**

- Competing risks are often present in analyses of a non-terminal event.
- The quantity of interest is usually the cumulative incidence function (CIF).
- Competing risks and right censoring are different: the event of interest cannot occur after the competing event has occurred.
- Treating competing events as right-censored observations leads to overestimation of the CIF.
- Always give the CIF for the competing risks as well as the CIF for the event of interest.
- If the competing events are not of interest, they can be grouped together as a single competing event.

### D. Recurrent events

Recurrent events arise when the event of interest can be experienced several times for each individual. In this case, a classical quantity of interest is the average number of recurrent events that a patient will experience up to a given time point, which is usually referred to as the cumulative mean number of recurrent events. Recurrent events occur when the event of interest is (for example) cancer recurrence, an infection, or hospital admission and when the objective is to estimate the average number of such events that a patient will experience up to any time point. Since censoring often occurs in this type of study, dedicated methods again have to be used to estimate such quantity of interest – typically, by appropriately estimating the hazard rate. In particular, ignoring censoring will clearly result in underestimation of the true number of recurrent events since censored patients will be followed-up on a shorter time period as compared to the situation where censoring did not occur. Furthermore, death is often observed as a competing event in medical studies (also called a terminal event in recurrent event studies) and must be accounted for; patients will not experience a recurrent event after death.

An estimator for the cumulative mean number of recurrent events was first developed simultaneously by Nelson and Aalen in the context of right-censoring; it is therefore referred to as the Nelson-Aalen estimator (20). The estimator was subsequently extended by Ghosh *et al.* to the case in which a terminal event is also present (21). Ghosh *et al.* also developed formulas for confidence intervals. It is important to stress that those formulas are very general and do not make any assumptions about the dependence structure of the recurrent event increments. In particular, they do not assume that recurrent events have independent increments, which is very often not the case in practice. More precisely, the recurrent events that may occur between any two time points are not assumed to be independent. In practice, this means that having already experienced one or more recurrent events may or may not influence the risk of further occurrences. This is a remarkable feature of the confidence interval formula because in practice, patients that have already experienced an event are often more likely to experience future occurrences (22).

We have developed code to generate recurrent events and have also implemented Ghosh *et al.*’s formulas for the estimator of the cumulative number of recurrent events in the presence of a terminal event (21). For the code, we refer the reader to the ***<u>Appendix</u>***. In order to implement the estimator, the dataset needs to be arranged in a start, stop structure (also called a counting process data structure). Each patient needs to have one line one line per recurrent event and one line for the censoring or terminal event time. On each lines, the start time is the occurrence of the previous recurrent event and the stop time is the occurrence of the next recurrent event. On the first line, the start time will be equal to the time when the patient enters the risk set (generally 0) and the stop time will be the censoring or terminal event time. This structure can also take into account left truncation: in such a case, the truncation time will be the start time of the first line. An example of data generated using this structure is given in **Table 2**.

**Table 2:** the first five rows of the simulated counting process database.

| Id | start | stop | status | terminal |
| --- | --- | --- | --- | --- |
| 1 | 0.00000 | 13.437412 | 1 | 0 |
| 1 | 13.43741 | 23.040031 | 1 | 0 |
| 1 | 23.04003 | 24.159902 | 1 | 0 |
| 1 | 24.15990 | 26.020856 | 0 | 1 |
| 2 | 0.00000 | 6.701158 | 0 | 0 |

Patient #1 developed an event three times (at the age of 13.4, 23 and 24.2 years) and died at the age of 26. Patient #2 did not experience any events and was censored at the age of 6.7 years.

Of note, the *survSplit* function from the *survival* library can also be used to create the counting process database.

The expected number of recurrent events is calculated using Ghosh *et al.*’s formula (21). We compared it with the naïve approach, which ignores censoring and the terminal event (***Figure 4***). The naïve estimator was implemented by simply counting the number of recurrent events that had occurred before a given time point, divided by the sample size. While the correct estimator only includes patients at risk of experiencing a recurrent event in the risk set, the naïve approach uses a fixed risk set that includes all the patients in the study. Since the risk set is too large in the naïve approach, the estimator underestimates the expected number of recurrent events. As recommended in the previous section, we advocate to also display the CIF of the terminal event because individuals at high risk of death will tend to experience fewer recurrent events. We computed it as one minus the Kaplan-Meier estimator (**Figure 4**, right-hand panel). As time goes on, the CIF moves closer to 1. This explains why the frequency of recurrent events appears to decrease slightly (on the left-hand panel); at late time points, the competing event is more likely to have occurred.

**Figure 4:**
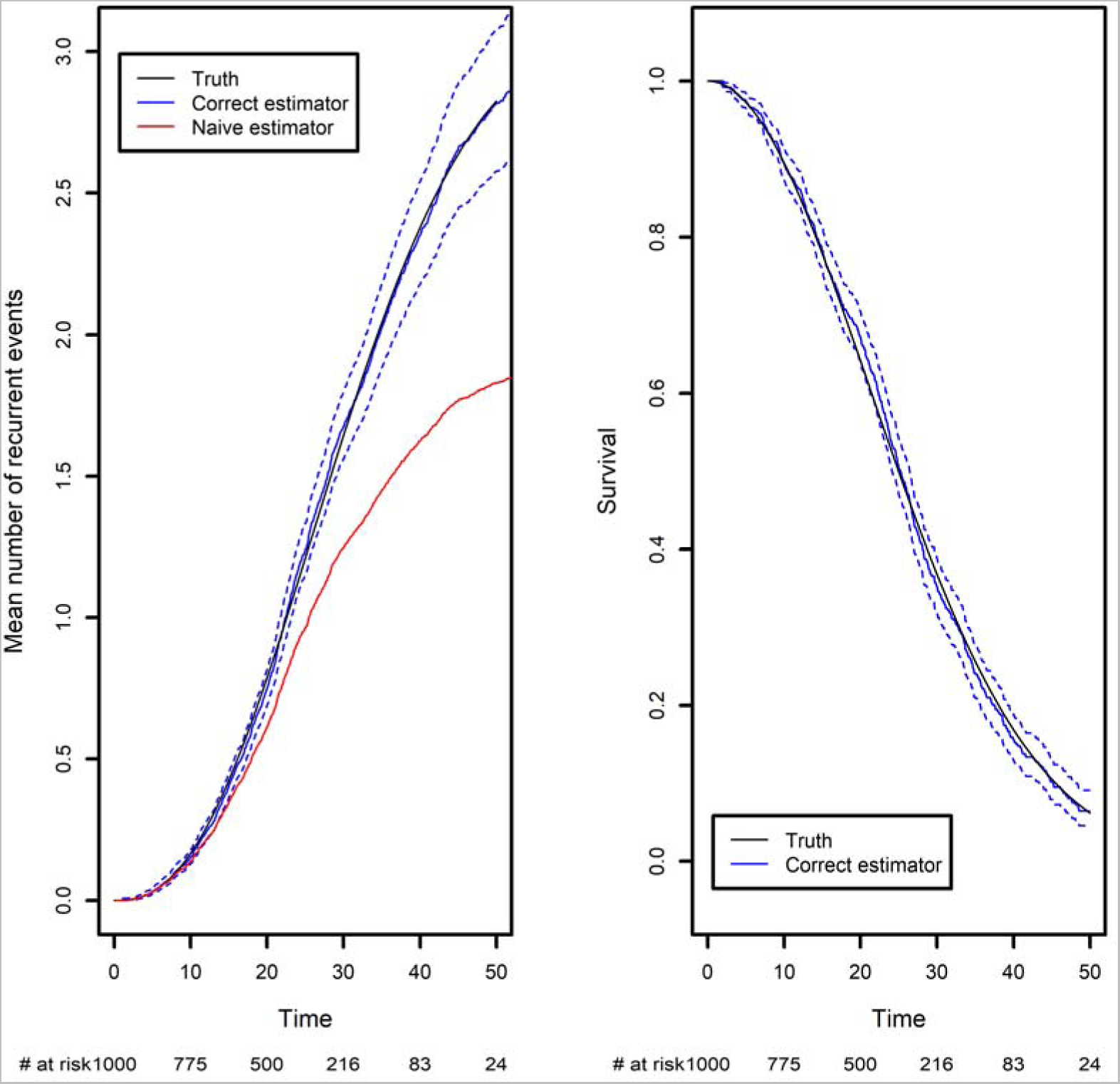
Comparison of the estimated mean number of recurrent events using censored data with competing events, and the naïve estimator that does not take into account censoring and the competing event. Left panel: the mean number of recurrent events. Right panel: the competing event (the terminal event).

Recurrent events often occur in medical registry dataset with often long follow-up periods and many repeated measurements of medical outcomes. Researchers are often not aware of the right method for handling recurrent events, and it is customary to analyze only the first event. This can lead to an important loss of information even though a recurrent analysis can be straightforwardly implemented using standard libraries for survival data. In order to compute the confidence intervals under the general dependence structure of the recurrent event increments, we implemented Ghosh *et al.’* s formula (21). Our code is available in the ***Appendix*** and can be applied to any recurrent event situation. At the time of writing, no packages were publicly available, and so we decided to implement the formula ourselves. Very recently, Klaus Holst and Thomas Scheike implemented a new function in the {*mets*} R package, which computes Ghosh *et al.’*s estimator. Both approaches give the same estimations.

#### TOOLBOX 5

**Recurrent events**

- Recurrent events occur when patients may experience the same event repeatedly over time.
- The cumulative mean number of recurrent events is an interesting summary measure of the frequency evolution of recurrent events over time.
- Competing risks often occur in recurrent event analyses.
- Ignoring right censoring will result in underestimation of the mean number of recurrent events.
- Treating competing risks as right-censored observations will lead to overestimation of recurrent events.

## 4. Use of the CEREDIH registry: a real case

PIDs are a very heterogeneous group of rare immune system diseases cause by defects in 485 genes (according to the latest international classification (6)). From a medical point of view, it is not usually relevant to analyze all the patients’ data together; usually, appropriate statistical analyses are conducted on subgroups of PID patients. Indeed, the data include patients suffering from very different diseases, such as T-cell deficiencies (mainly SCIDs and CIDs), B-cell deficiencies, and innate immunodeficiencies (***Figure 5***).

**Figure 5:**
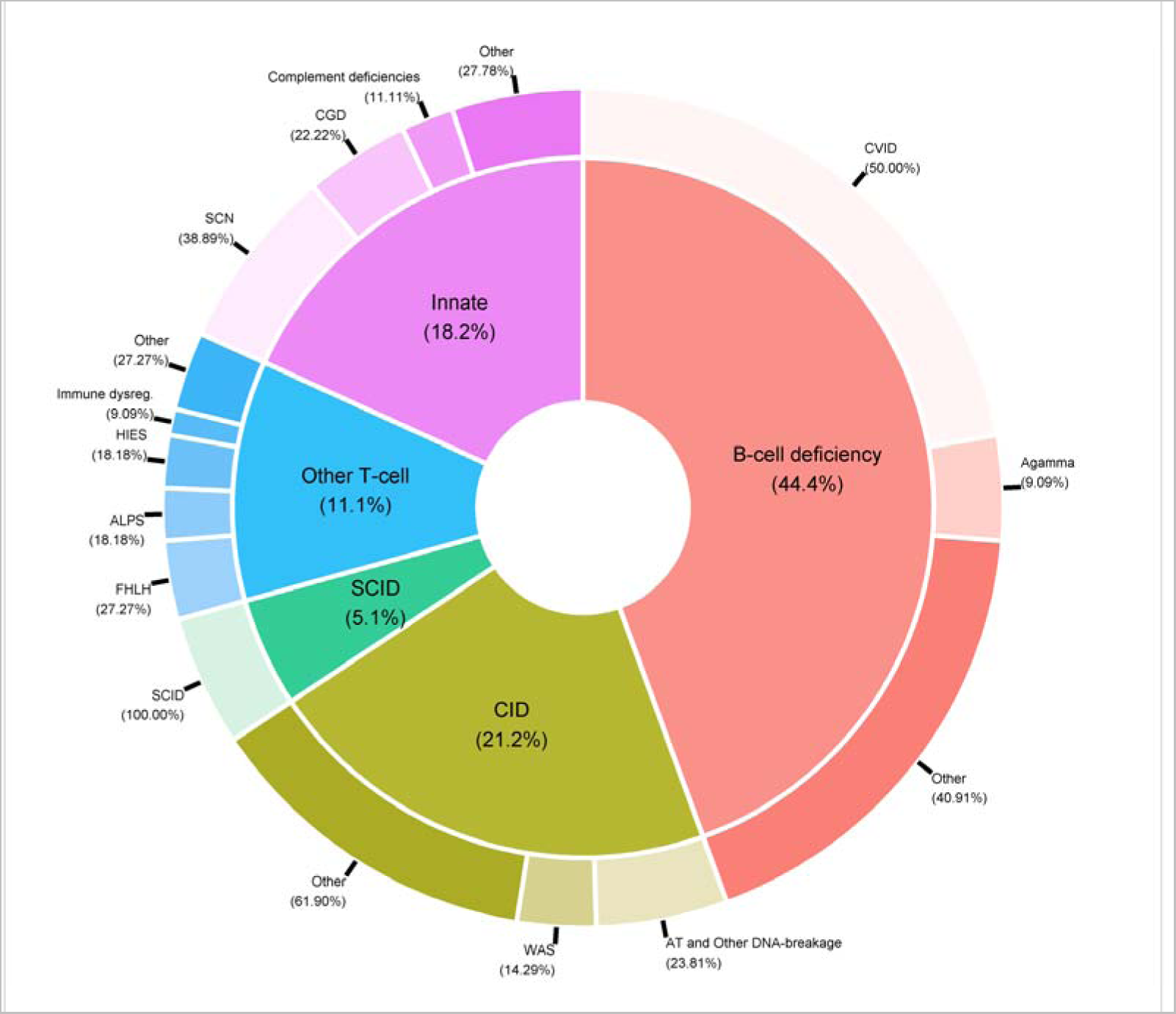
Distribution of PID categories in the CEREDIH registry (n=7,753 patients) <u>as of June</u> 22^nd^, 2022

The data in the CEREDIH registry are typically right-censored because they are collected in real time; as a result, most patients are alive at the time of registration. The data are also retrospective in the sense that deceased patients can also be registered if they were diagnosed with a PID before death. In fact, the diagnosis of PID is a requirement for registration. This means that PID patients who die before being diagnosed are not included in the registry. This is a typical example of left truncation when studying the patient’s age at death and must be taken into account appropriately in the statistical analysis.

When dealing with time-to-event data, one must define the starting point for the follow-up of each patient. Ideally, the choice of the starting point is guided by medical considerations, i.e. the time that makes the most sense for the patient. For the CEREDIH registry, one possibility is to set the “start of follow-up” at the date of the clinical diagnosis. In that case, the time-to-event variable will be the time elapsed since diagnosis. Another possibility is to set the “start of follow-up” at the date of birth, in which case the time-to-event variable of interest will be age (i.e. age when the event of interest occurred). The latter option makes more sense from a medical point of view because PIDs are genetic diseases; even though a patient might be diagnosed at a later age, the disease might have affected him/her since birth or at least for some time before the diagnosis. As mentioned above, however, the date of diagnosis might be a truncation variable. In contrast, setting the start date to the date of diagnosis avoids the left truncation issue.

For the sake of clarity, we first show the Kaplan-Meier analysis of the time to death of patients with CVID when the starting point is the date of diagnosis. Patients with CVID are diagnosed at different ages, ranging from early childhood to late adulthood. Our analyses were further stratified with respect to five age classes. When patients are diagnosed at a later age (40+), the risk of death differs significantly (**Figure 6a**). Twenty years after diagnosis, 45% of the patients diagnosed after the age of 40 were dead. For patients diagnosed between 0-4, 5-9, 10-19, and 20-39 years of age, the death rate was 8%, 4%, 15%, and 9%, respectively. However, the patients diagnosed earlier in life are more prone to die for a reason unrelated to their PID; this highlights the limitations of this method and its interpretation. The same analysis was conducted for six different PIDs but with birth as the start date (***Figure 6b***). This time, the left truncation induced by the date of diagnosis was taken into account by applying the above-described methodology. We can see that CVID and non-CVID B-cell deficiency patients have similar survival curves and have a better prognosis than patients with the other diseases. At 40 years of age, for example, 12% of the CVID patients and 13% of the non-CVID patients are estimated to have died. Patients with an innate immunodeficiency tend to have a higher survival that patients with a CID: the probabilities of dying before 20 years of age are estimated to 25% for patients with an innate immunodeficiency and to 45% for patients with a CID. A high proportion of patients with SCID are estimated to die at a young age: the probability of dying in the first two years is estimated 44%. For patients who have survived, the probability of death is low. In the following examples, we will always use birth as the start date.

**Figure 6:**
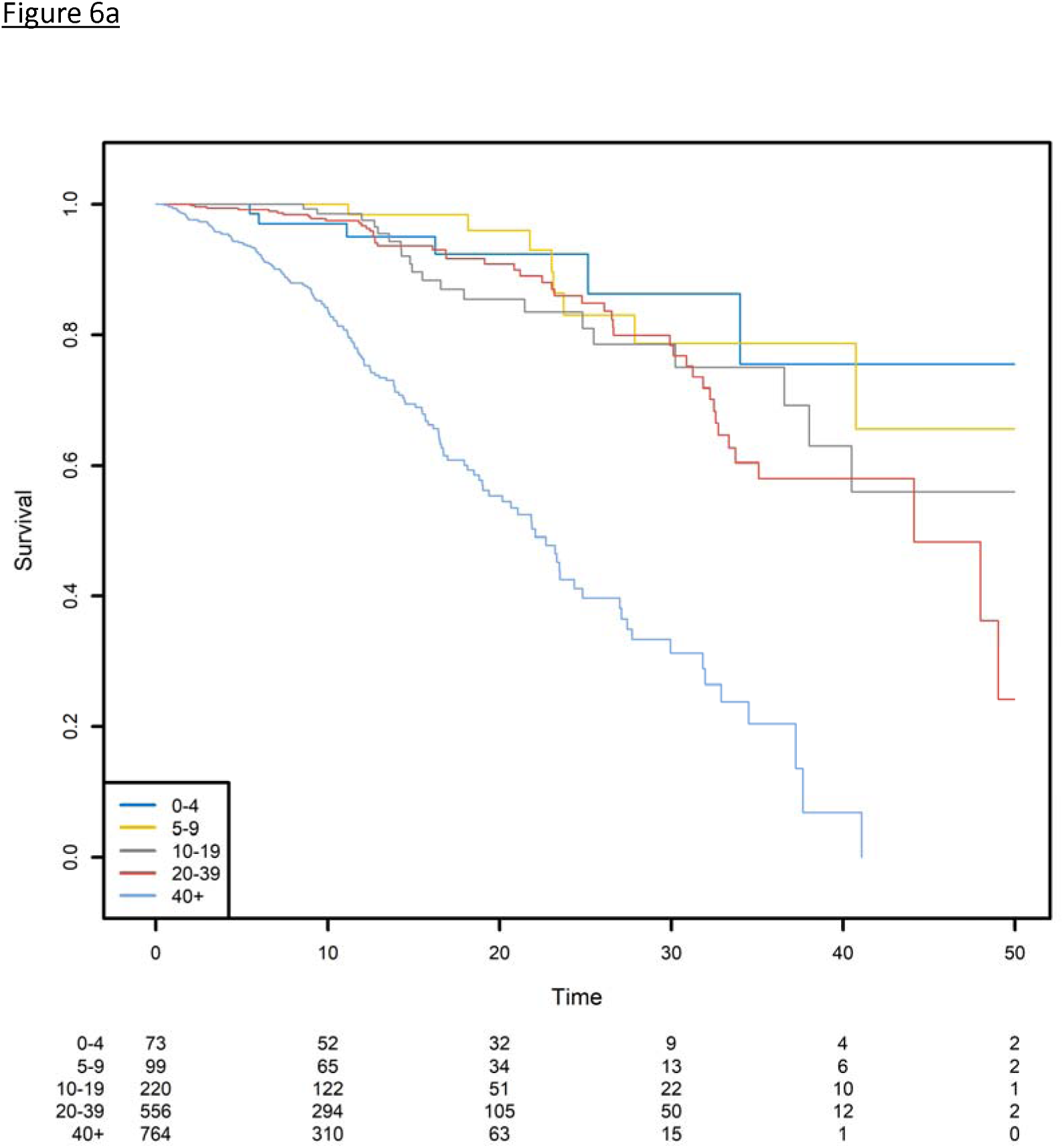
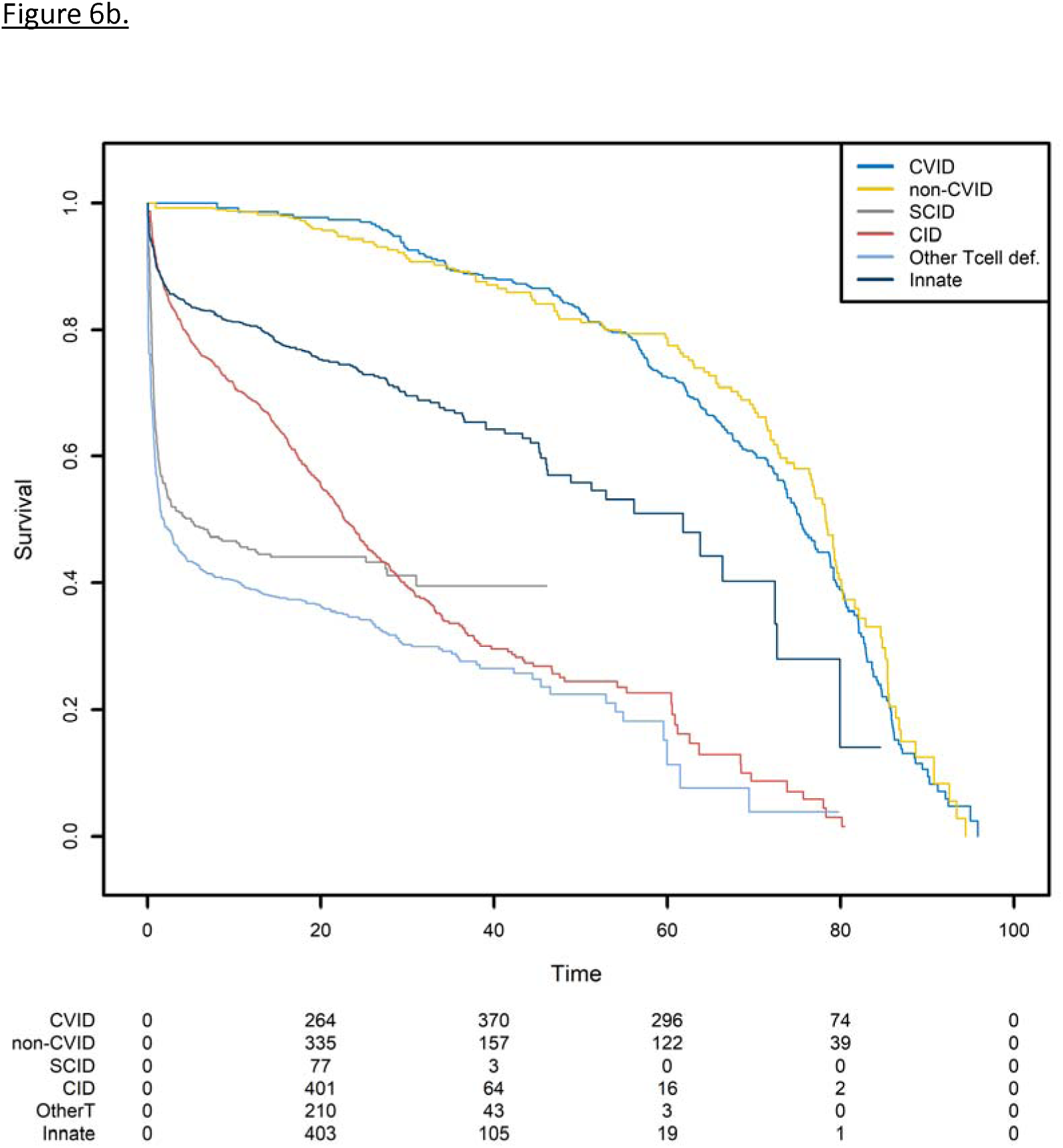
Survival probabilities with different starting points. (a) Post-diagnosis survival for patients with CVID, by age at diagnosis. (b) Overall survival from birth, by PID category.

Not taking left-truncation into account in registered patients with CVID leads to significant overestimation of the probability of survival (***Figure 7***). By using the correct methodology, we estimate that 30% of patients will die before the age of 62.3 years. When the naïve estimator is used, the equivalent age is 75.0. This comparison highlights the potential consequences for public health at the population level.

**Figure 7:**
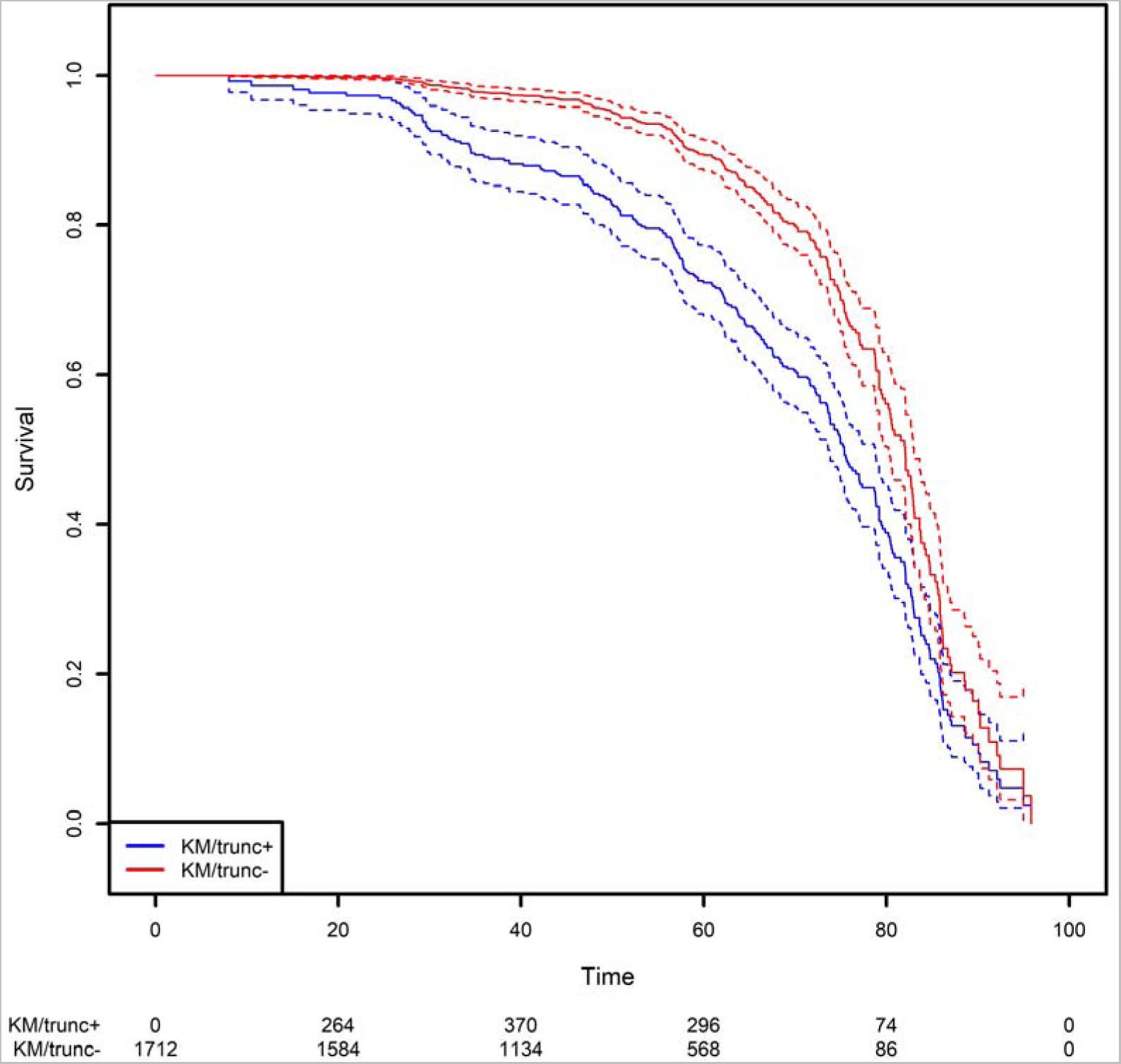
Comparison of the estimation of the survival function on the CVID population using the Kaplan-Meier estimator that takes left truncation into account and the KM estimator that ignores left truncation.

We next studied the first occurrence of cancer in patients with PID. Here, the event of interest is cancer, and so death is an obvious competing risk that needs to be handled properly. Since we are interested in the occurrence of cancers associated with PIDs, we also considered all curative therapies (HSCT, gene therapy, and thymus transplantation) as competing risks. These curative therapies can be treated as a single composite event recorded at the age when the patient first encounters a competing risk. We computed the CIF for cancer in the six subgroups of PID patients and the CIF for the composite competing risk (***Figure 8***).

**Figure 8:**
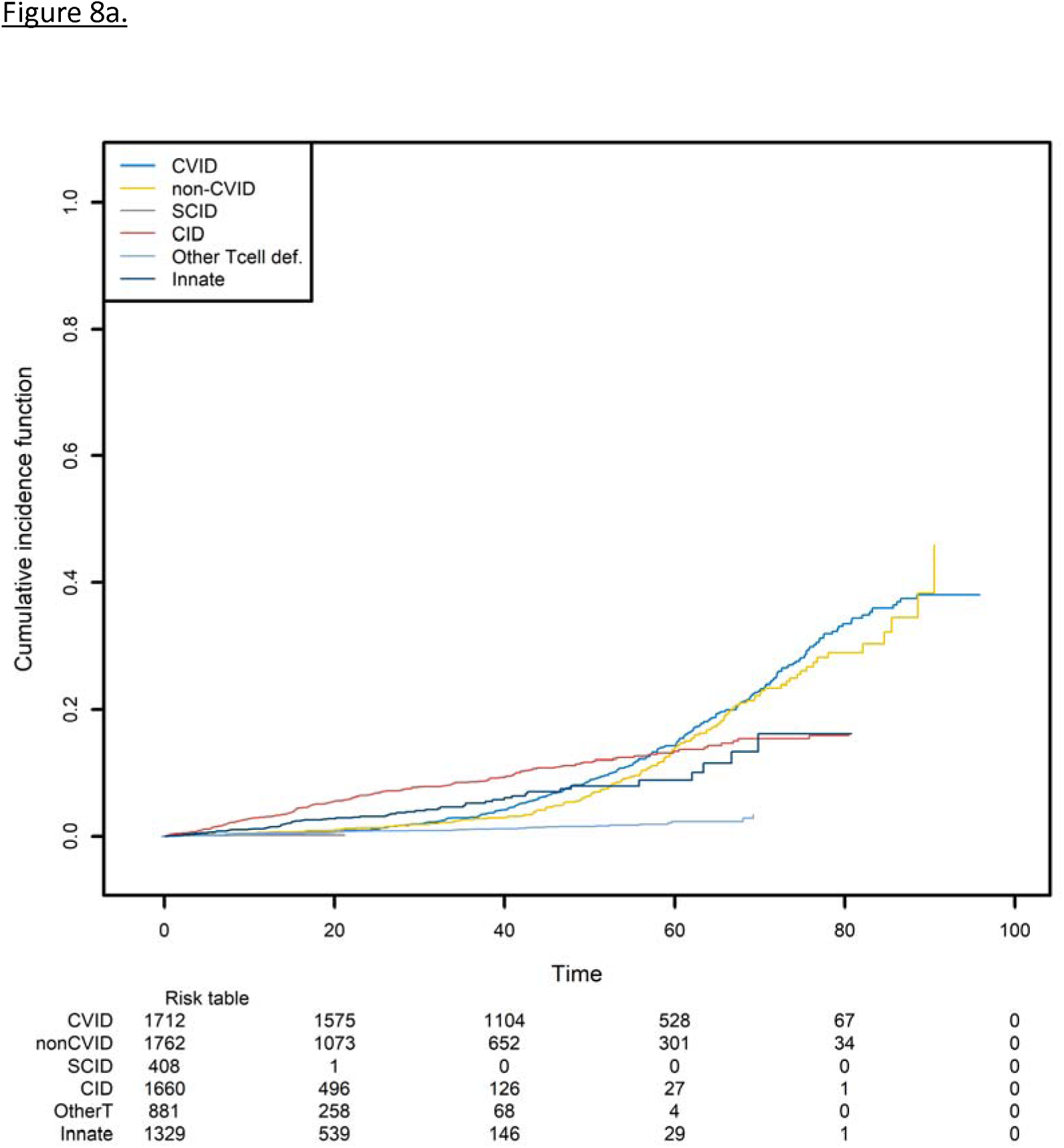
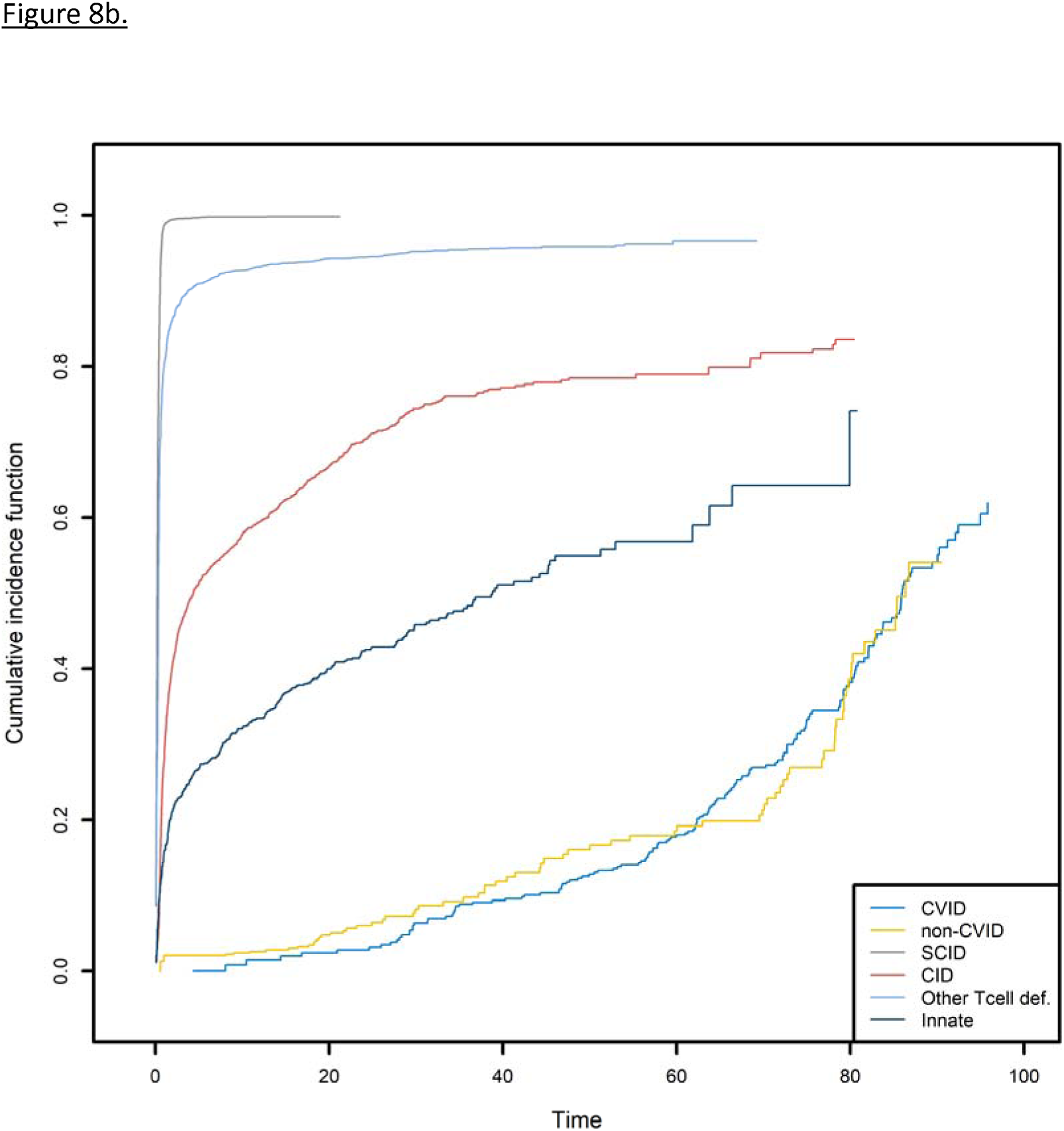
The probabilities of developing a first cancer and its competing event, according to the main PID categories. (a) the probability of developing a first cancer, according to the main PID categories. (b) the probability of death or receiving curative therapy (the competing risk), whichever comes first and according to the main PID categories.

CID patients are more likely to have experienced a first cancer before the age of 55 (***Figure 8a***). For the patients that are still alive at that age and have not yet experienced cancer or a curative therapy, the CVID and non-CVID patients are the most at risk of developing cancer. Clearly, these findings are strongly linked to those shown in ***Figure 8b***. Patients with SCID can undergo curative therapy or die very soon after they are born and so are no longer at risk of developing cancer. In contrast, the patients with a CVID or non-CVID B-cell deficiency have a much lower risk of death or curative therapy than the other patient groups; this is because of their greater risk of cancer at older ages.

Lastly, we analyzed the recurrences of cancer, autoimmune disease episodes, and inflammatory events on the CID patient group (**Figure 9**). Any of these three types of event is defined as a recurrent event. We sought to estimate the mean number of such events having occurred before any time point. As in the previous analysis, curative therapy and death are considered to be a composite competing event. The mean number of recurrences is low and increased slightly over time to a value of 0.74 before the age of 80. Again, this value must be compared with the risk of experiencing death or a curative therapy over time, which is high for these patients (0.98 before the age of 80).

**Figure 9:**
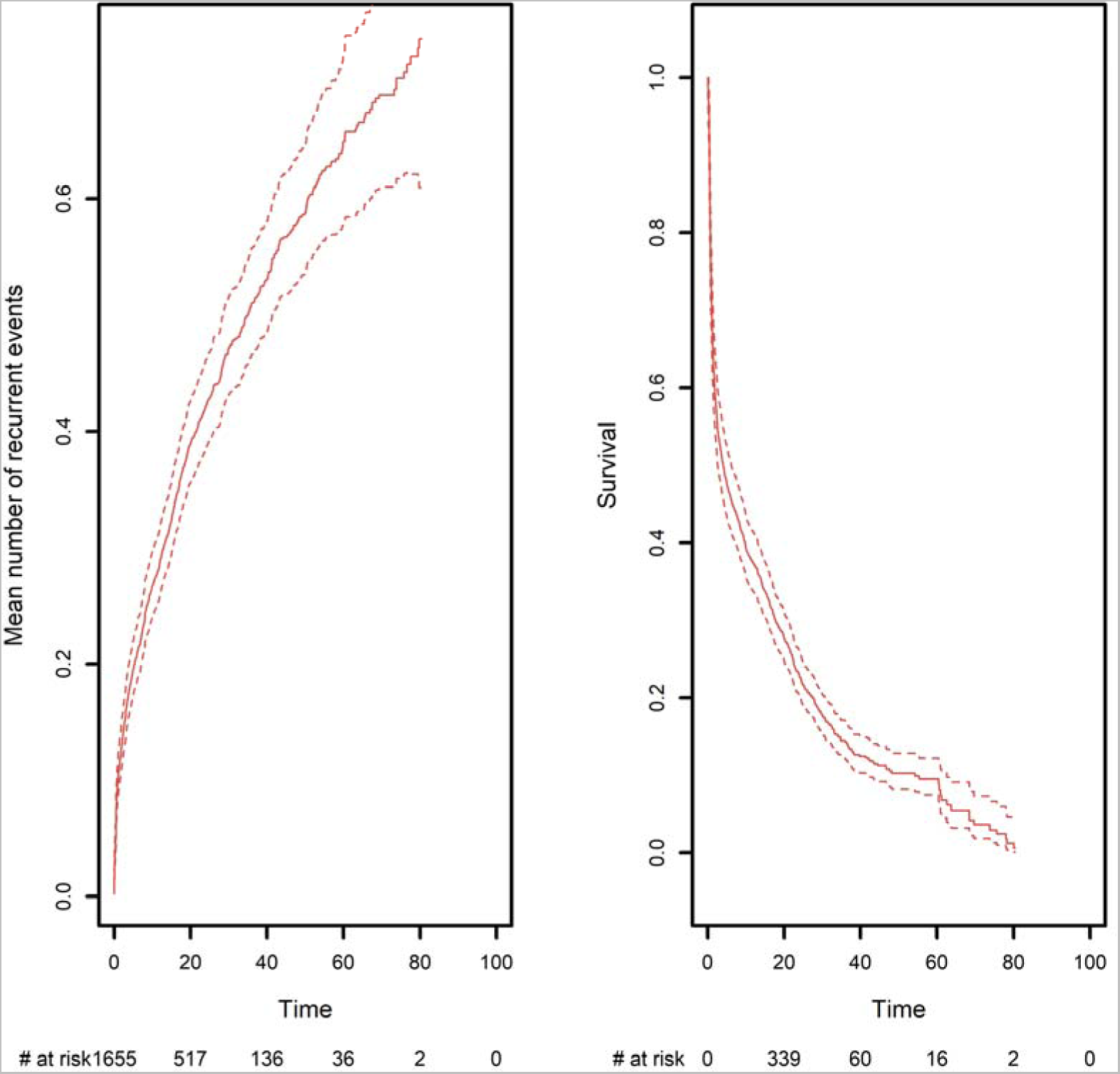
(a) the mean number of recurrent malignant, autoimmune, and inflammatory events, taking death and curative therapies as competing risks. (b) the competing event survival function, where the competing event is defined as death or curative therapy (whichever comes first).

## 4. DISCUSSION

In the present article, we discussed dedicated statistical methods for analyzing time-to-event data in registries. Those types of data have the particularity that they are not completely observed, and various approaches must be used to avoid biased estimations. In particular, we discussed how to take into account right-censoring, left truncation, and competing events. We also considered recurrent event situations, in which individuals can experience the event of interest several times. If those particular mechanisms in the collection of the data are not properly taken into account, the application of standard methods for completely observed data will give rise to systematic biases. This fact was highlighted in a simulation study in which the true mechanism that generated the data was known in advance; and it was therefore possible to compare various methods with the truth. Lastly, we used the methods presented here to analyze data from the CEREDIH registry.

In order to avoid bias in the analysis of time-to-event data, we refer to the three pillars stated by Andersen and Keiding (23). “First, do not condition on future”. In other words, no estimation should be carried out that uses events that will occur in the future. For example, when one wants to compute an estimation on the age scale, one should check whether all the patients have been followed up from birth. This is typically not the case when age is the time scale, and the data will probably suffer from delayed entry (i.e. left truncation). In the CEREDIH data, for instance, left truncation means that patients are included in the study because it is known that they will be diagnosed at some time in the future. Nevertheless, we have seen in this paper how left truncation can be taken into account by modifying the risk set using the age at diagnosis. The second principle is “do not regard individuals at risk after they have died”, and the third is “stick to this world.” In the present article, these last two points apply to competing risks. Censoring a patient at the time of his/her death implies that he/she will experience the event of interest postmortem, which is impossible in the real world.

Left truncation is a specific example of immortal time bias. Immortal time bias occurs when an individual is incorrectly considered to be at risk, i.e. during a period of time when they cannot experience the event of interest (24). As an illustration of immortal time bias, let us consider a cancer study in which the objective is to compare the risk of death for a group of patients with cancer and a group of patients without cancer. If the time scale is age, then cancer status will be a truncation variable. Furthermore, the cancer and non-cancer groups are not well defined because it is not possible to know in advance (i.e. at birth) whether the individual will remain cancer-free or will develop cancer at a later time. If one performs a survival analysis by defining the two groups (cancer and non-cancer) in advance using the Kaplan-Meier estimator, then the survival curve of the non-cancer group will be strongly biased downwards; the risk set will not include any of the patients in the cancer group, even though many of these patients will not have yet developed a cancer at some specific times and should therefore be included in the risk set. Since the non-cancer risk set will be much smaller than it should be at early time points, the corresponding survival curve will indicate that the prognosis for the non-cancer group is worse than it truly is. In contrast, the fact that the cancer group is defined prior to the onset of cancer introduces selection bias. As a result, the comparison of the two curves will largely attenuate the effect of cancer on death. It is important to note that with age as the time scale, the cancer group does not correspond to a real situation. Individuals are not born with a cancer status, and cancer may or may not occur during the lifetime of a patient. When cancer does occur, the individual is no longer at risk of developing cancer but might have an elevated risk of death. In fact, this is a multi- state situation in which different events with different risks must be taken into account in the survival analysis.

It was not possible for us to cover all the types of incomplete observation that can arise in analyses of time-to-event data. Furthermore, we did not discuss how to analyze the effect of covariates on a time-to-event response variable through regression modelling. In the context of right-censoring and/or left truncation, this is usually performed with the Cox model (16). The Cox model can also be applied to competing events and recurrent events, notably via the survival package in R. Lastly, MSMs constitute a major topic of interest but are not covered here (17). This situation arises when (i) an individual can experience different events (referred to as states) during his/her lifetime and (ii) the risk of experiencing any of these events varies. This is a natural extension of the competing risk situation; instead of studying only two possible events (one of which is terminal), one looks at multiple events between which transitions may or may not be allowed in the model. In the CEREDIH registry data, for instance, patients may experience various events: severe infections, cancer, auto-immune disease episodes, death, etc. By using MSMs, one can describe all the different states associated with the disease and the changes from one state to another. This approach can also be incorporated into a regression model in which covariates affect some states and not others. These regression models can be implemented using the *{msm}* or *{mstate}* packages in R.

In this article, we presented various methods and highlighted a number of pitfalls in the analysis of time-to-event data. As a consequence, we strongly encourage medical researchers who study time-to-event data to collaborate closely with statisticians. Firstly, registry data (especially rare disease registries) are essential for understanding a disease (10). Secondly, funds for rare disease research are often limited, and it is therefore crucial to use appropriate statistical methods and derive correct conclusions. Robust, high-quality health data are critical for (i) enhancing healthcare delivery, medical R&D, and our knowledge of disease, (ii) supporting policy and regulatory decisions, and (iii) ultimately benefiting patients in particular and society more widely. Data can change lives by speeding up diagnosis, improving patient care, and fostering the development of new treatments. In rare diseases like PIDs, health data is even more vital for the provision of more effective, high-quality, safe and personalized care. Worldwide, efforts are growing to strengthen the collection and use of data through patient registries and the shaping of collaborative health data ecosystems.

## Data Availability

The data are not publicly available

## Standfirst

Based on both the use of real-world data and a simulation approach, this tutorial article looks at how overlooking right censoring, left truncation, competing events and recurrent event methods in analyses of time-to-event data can lead to suboptimal or biased estimations. The simulation approach enables a comparison with the truth and thus concrete estimations of bias for methods that (i) do not consider right censoring (leading to underestimation of survival), (ii) do not consider left truncation (leading to overestimation of survival), (iii) treat competing risks as right-censoring (leading to overestimation of survival), and (iv) consider only the first of a series of recurrent events for a given individual.

## Contributors and sources

In this tutorial article, we look at how overlooking right censoring, left truncation, competing events and recurrent event methods in analyses of time-to-event data can lead to suboptimal or biased estimations.

The present work is novel because it uses both real-world data and a simulation approach. This enables comparisons with the truth and thus the provision of concrete estimations of the biases associated with various methods.

Mickaël ALLIGON is a junior statistician at the French National Registry for Primary Immunodeficiencies (CEREDIH). He trained under the supervision of CEREDIH director Nizar MAHLAOUI MD, PhD (a clinician and epidemiologist with 18 years of experience) and Olivier BOUAZIZ PhD (a senior statistician in the Laboratory of Applied Mathematics in Health Sciences, with great expertise in survival analyses). All the authors contributed equally to analyzing the data and drafting the manuscript. MA and OB performed the statistical analyses.

OB is the guarantor of the article. The CEREDIH registry was the source of the real-world data. This research did not receive any specific funding from agencies or organizations in the public, commercial, or not-for-profit sectors.

## ABBREVIATIONS

CID: combined immunodeficiency
CIF: cumulative incidence function
CVID: common variable immunodeficiency
HSCT: hematopoietic stem cell transplantation
MSM: multistate model
PID: primary immunodeficiency
SCID: severe combined immunodeficiency

## List of Supplementary Material

Supplementary data 1: True cumulative incidence function in a competing risk situation, using two Weibull distributions for the simulation

The two competing risks are simulated by two Weibull distributions. k_1_ and k_2_ are shape parameters, and λ_1_ and λ_2_ are scale parameters.

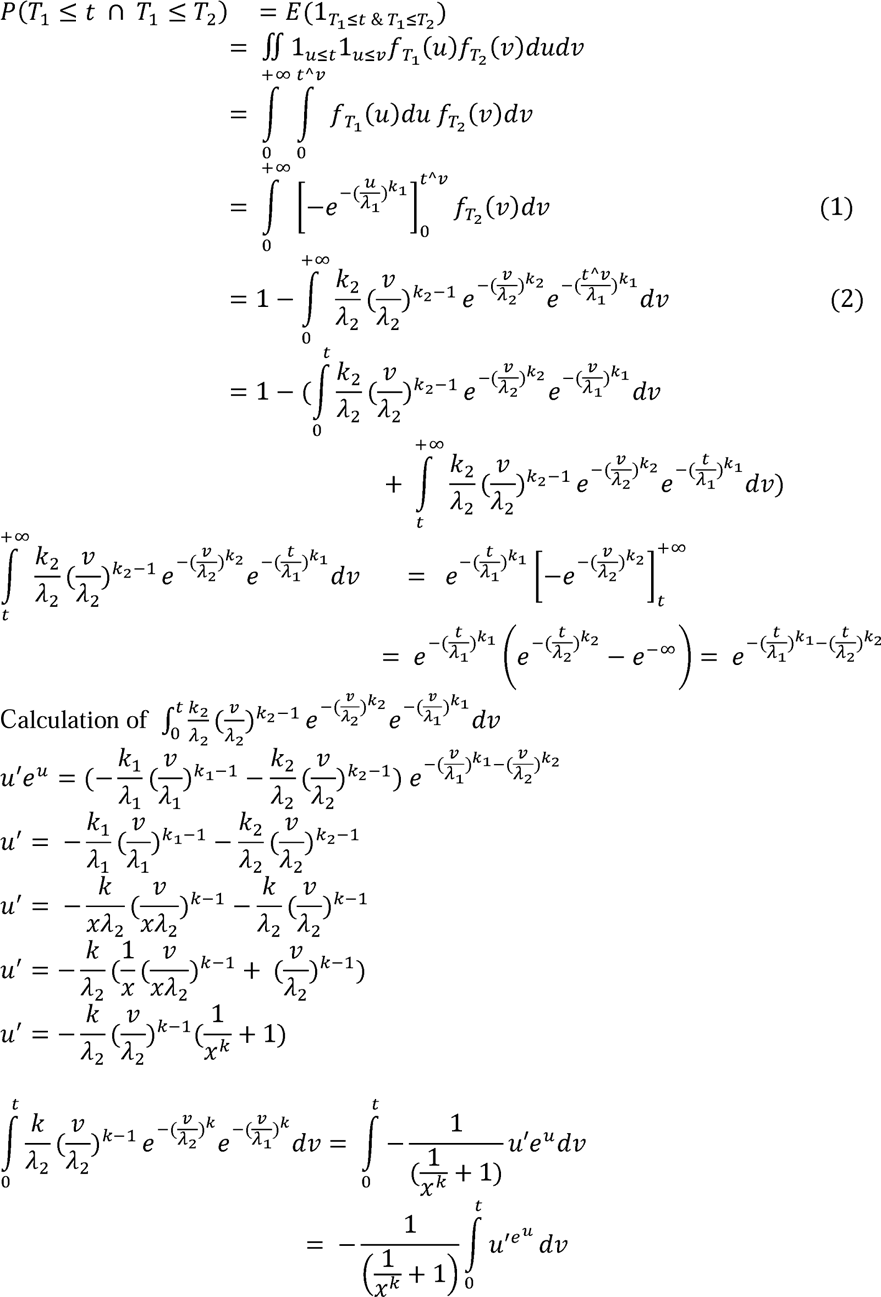

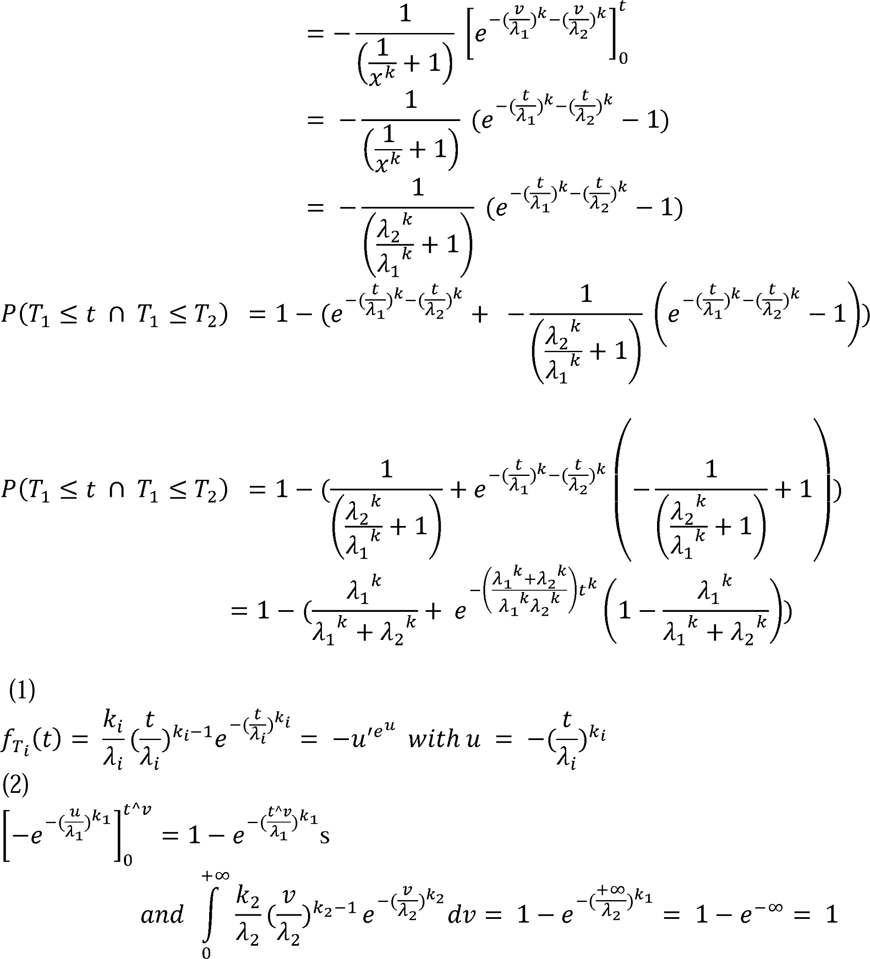

## Appendix

https://github.com/Malligon/Pitfalls-in-Time-to-Event-Analysis-for-Registry-Data

